# Surgical Site Infections in Tanzania: A Five-Year Trend and Variation Analysis of National Surveillance Data, 2021-2025

**DOI:** 10.64898/2026.09.09.26362600

**Authors:** Cesilia Charles, Jonhas Masatu, Furahini Mbise, Omary Nassoro, Lackson David, Laura Marandu, Calvin Andrea, Melkizedeki Abdulahi, Upendo Msanjila, Nuru Beda, Christina Mmasa, Adam Kamese, Elford Mukerebe, Erick Kinyenje, Radenta Bahegwa, Siril Kullaya, Edgar Lusaya, Joseph C Hokororo, Eliudi S. Eliakimu

## Abstract

**Background:** Surgical site infections (SSIs) are an alarming threat in the surgical field worldwide. They are among the most prevalent hospital-acquired infections (HAIs) and contribute to antibiotic resistance in resource-limited countries, including Tanzania, where data remain scarce. This study aimed to ascertain the five-year trend of SSI in Tanzanian health facilities.

**Methods:** A retrospective descriptive cross-sectional study was conducted using SSI data reported in the District Health Information System-2 (DHIS2) between 1^st^ October 2021 and 30^th^ December 2025. Data from 138 health facilities were extracted, cleaned, and analysed in R (version 4.5.0). Trends in SSI rates were assessed using Ordinary Least Squares (OLS) regression, with significant variations measured at ≤ 0.05.

**Results:** The 138 facilities reported 419,412 surgical procedures during the study period. Most facilities were primary-level (96, 69.6%) and government-owned (119, 86.2%). The number of reporting facilities increased from 17 in 2021 to 138 in 2025, with an increase in surgical procedures from 9,616 to 127,039. Overall SSI rates declined from 3.1% in 2021 to 2.4% in 2025. Primary-level facilities had the highest overall SSI rate (5.4%), declining from 4.7% to 4.0%. In contrast, SSI following section increased from 1.8 % to 2.0%. Organ/space SSI showed a statistically significant declining trend from 1.2% to 0.3% (slope = -0.23%/year; 95% CI: -0.34 to -0.12; p = 0.006). The trends in overall SSI rates by level of health facility and ownership were not statistically significant

**Conclusion:** Between 2021 and 2025, overall SSI rates decreased despite a substantial increase in surgical volume, suggesting improvement in IPC. While an organ/space SSI significantly decreased, SSI rates due to caesarean section persistently increased, with higher rates reported in primary healthcare facilities, highlighting the need for strengthened perioperative IPC measures in obstetric care and SSI surveillance across primary-level facilities.

## Introduction

Globally, SSIs remain a significant public health challenge and a major postoperative complication worldwide, accounting for one-fifth of all HAIs ^1^. It is among the most commonly reported HAIs occurring within 30 days after a surgical procedure or up to 90 days for implants. Based on the location and depth of the infection, SSIs are classified as superficial (infection involves only skin and subcutaneous tissues of the incision), deep (infections that involve deep soft tissues of the incision, for example, fascial and muscle layers), and organ/space (infections that involve the organ/space tissues deeper than the fascia/muscle)^2^.

The global burden of SSI remains substantially high, with lower- and middle-income countries (LMICs) accounting for up to 20%, a higher proportion compared with 2-5% in high-income countries ^3,4^. In Sub-Saharan Africa (SSA), pooled estimates suggest SSI prevalence exceeding 14% with wide variation across the region and facility levels^5^. In Tanzania, hospital-based studies report SSI rates to be between 10% and 48%, reflecting systemic challenges, limited infection prevention and control (IPC) supplies, poor surgical practices, and inconsistent monitoring and evaluation systems ^6^. Even though the SSIs are preventable, they are associated with negative health outcomes such as increased antimicrobial use, increased morbidity, mortality rates, and readmission^7^. Beyond these health risks, SSIs pose substantial economic challenges for both the individual and the healthcare system due to prolonged hospital stays by 10 days and increased direct medical costs by 300% to 400% ^8^. On the other hand, SSIs compromise surgical safety and undermine progress towards universal health coverage ^9^.

In response to SSIs, multiple interventions have been implemented globally and nationally, including the distribution of evidence-based guidelines produced by the World Health Organisation (WHO) on SSI prevention, which address preoperative antibiotic prophylaxis, surgical hand preparation, sterile techniques, and postoperative wound care^10^. Tanzania has adopted IPC guidelines and strengthened quality improvement initiatives through programs such as Routine IPC assessment and mentorship through standard-based management and recognition (SBM-R), infection prevention and control assessment framework (IPCAF), Clinical Audit, and antimicrobial stewardship frameworks ^11–13^. Additionally, regional and district health facilities in some regions have implemented routine SSI surveillance and reporting systems^14^.

Despite these ongoing efforts, the Tanzania Ministry of Health and stakeholders have integrated the SSI indicator into District Health Information System 2 (DHIS2), aiming to establish national, regional, council, and health facility data to inform decision-making, policy formulation, and improve service delivery, hence reducing SSI. However, since integration, there is a limited number of national SSI data analyses that have been done to assess the trend and variation of this indicator across the regions and health facilities. The few existing studies were conducted in a single centre using primary data and cross-sectional designs or limited to short timeframes ^6,15–18^, thereby restricting the ability to assess longitudinal trends and regional heterogeneity across the country. The analysis of routine health facility (DHIS2) data plays a vital role in monitoring service delivery, particularly for tracking the SSI burden in countries with high SSI mortality rates, such as Tanzania ^20^. This monitoring is crucial, especially as Tanzania is currently off track to achieve Sustainable Development Goal (SDG) 3.8. Therefore, this study evaluated trends and variations in SSI across Tanzanian health facilities from 2021 to 2025 to inform evidence-based policy, optimize resource allocation, and strengthen surgical quality.

## Materials and Methods

### Study design and setting

This descriptive cross-sectional study was conducted in Tanzania to determine the national trends and variation of SSIs across Tanzanian health facilities using the DHIS2 data from 1^st^ October 2021 to 30^th^ December 2025. Tanzania, with a population of 61.7 million and a population growth rate of 3.2, has been classified as a lower-middle-income country since 2020 ^21^. The country has 26 administrative regions, 184 Local Government Authorities (LGAs), also referred to as Councils ^22^ and the country’s healthcare delivery is based on a three-tier system as follows:

- Primary level includes i) community services, offering health promotion, disease prevention, and some curative services provided by the community health care workers (CHWs), ii) dispensary – is the lowest facility level serving one or a few villages or a ward and primarily focuses on providing outpatient care, iii) health centre-offers a higher level of care compared to the dispensary and serves a larger population, is also required to provide inpatients and receives referrals from nearest dispensaries and iv) hospital at a district council-serves as the primary referral within the district and provides more comprehensive healthcare services ^23^.
- Level 2(secondary) includes referral hospitals, regional levels, among other services, provide the first level of specialized health services and serve as referral centres for patients requiring advanced medical services beyond the capabilities of primary healthcare facilities.
- Level 3 (tertiary) includes referral hospitals at the zonal level, specialized hospitals, and national hospitals, which provide high-tech, specialized, and complex medical care and serve as teaching hospitals for medical, paramedical, and nursing training. These facilities are equipped to handle advanced medical care ^24^.

### Study population

The study included 148 health facilities whose healthcare workers (HCWs) have been trained on the SSI case definition, case follow-up, and how to report cases in the DHIS 2 platform monthly. We excluded eligible health facilities that had incomplete data of more than 5% and those with missing data elements on SSI. Thus, the final analysis included 138 health facilities that had fulfilled the eligibility criteria. (Fig 1)

**Fig 1:**
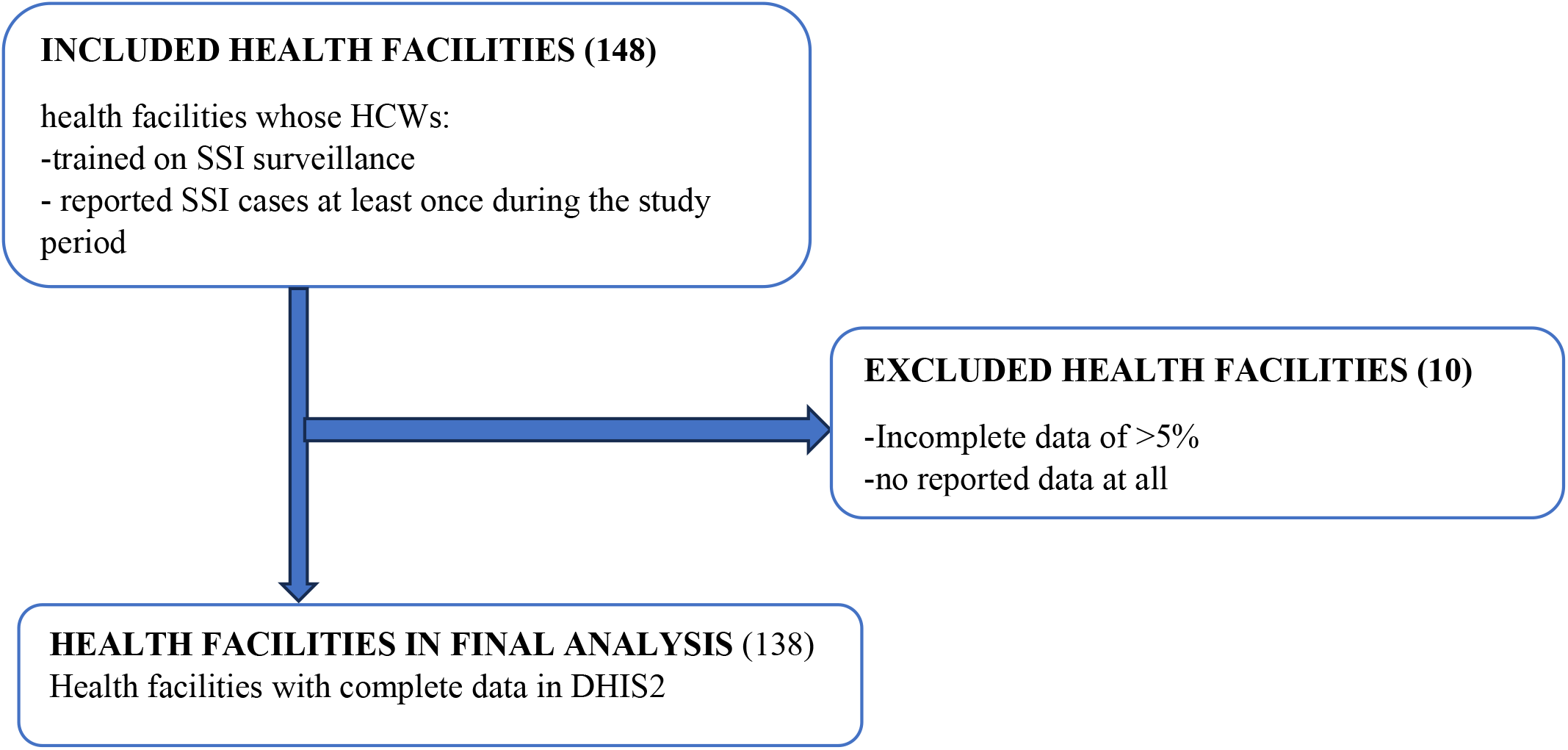
Enrolment of the study participants during the study period of 2021-2025

### Data source

The study was based on secondary analysis of routine health facility data available through the DHIS2 database from 1^st^ October 2021 to 30^th^ December 2025. DHIS2 is a platform recommended by the WHO for managing health information in low-resource countries, including Tanzania. It has been introduced into the Tanzanian health management information system to facilitate monthly routine evaluation and monitoring of health services delivery. In this system, the initial recording of events is conducted using surveillance forms and paper-based registers at the facility level, where HCWs document primary data from various wards. For some dispensaries, health centres, district hospitals, regional and tertiary hospitals with HMIS departments, data is entered directly into DHIS2 within the health facilities, while some dispensaries and other lower health facilities, in the absence of a dedicated DHIS2 focal person, submit their monthly paper-based reports to the district information system focal person for entering them in DHIS2. Non-public hospitals (private and faith-based) are also required to submit these reports to DHIS2. (Fig 2).

**Fig 2:**
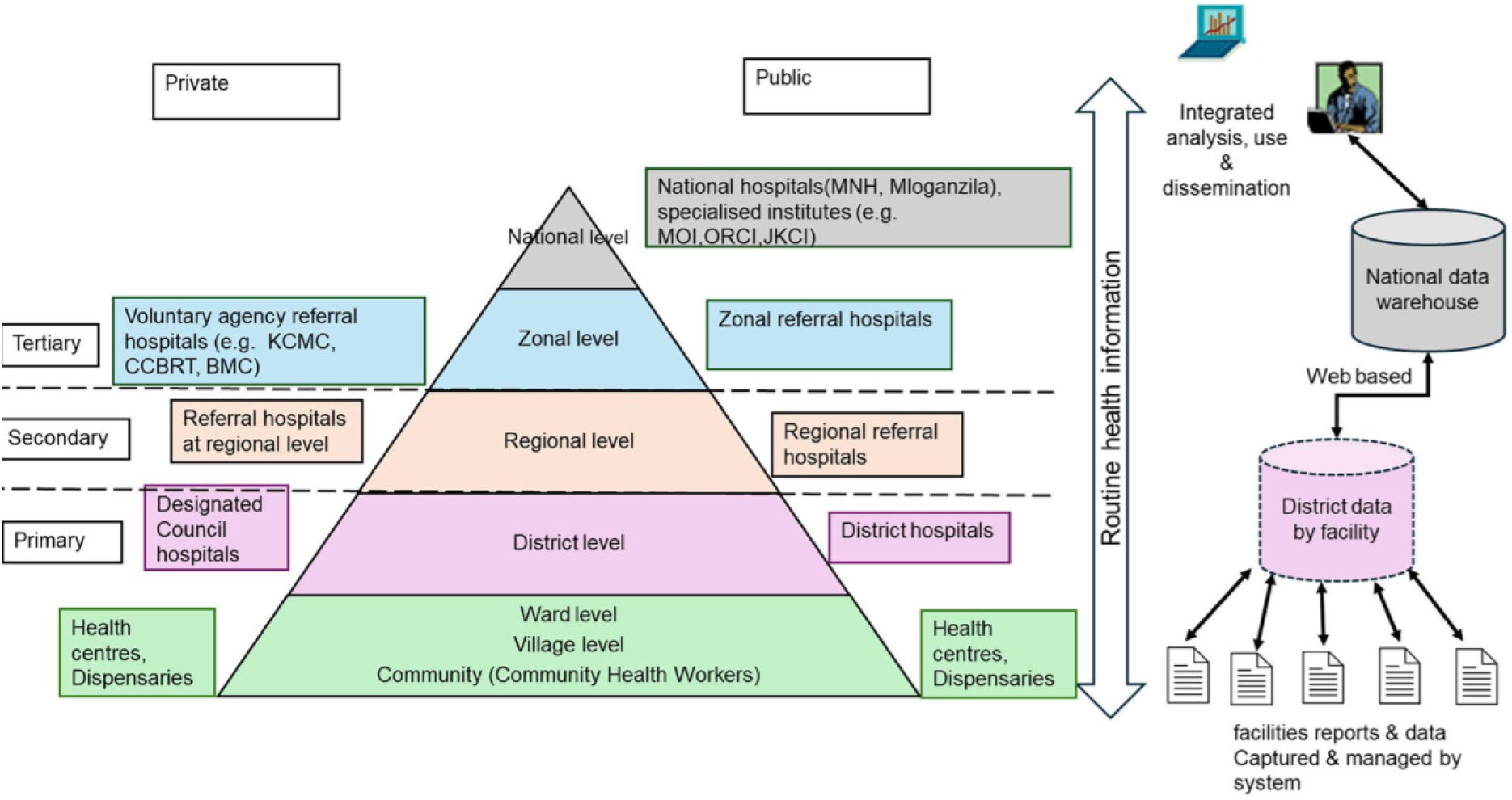
Tanzania Health System and Process of Data Collection in DHIS2 ^25^

The SSI indicators are captured in various health facility registers, including surgical wards, minor theatres, postnatal wards, minor theatres, and Reproductive and Child Health (RCH), before being entered into surveillance forms (denominator and numerator forms) then the IPC daily register. The SSI data are tallied in the facility IPC registers and then entered into DHIS2.

### Data Quality

Data quality for the SSI indicator was ensured by collecting surgical procedures and SSI records from health facilities whose HCWs have been trained in the SSI case definition and in routinely reporting their data in DHIS2. Additionally, data were extracted from DHIS2 (the national health information system) with embedded validation rules, and, during analysis, data governance practices were applied to ensure the data remained an exact reflection of reported values, with no transformations that could introduce bias.

### Data Management and Analysis Plan

The extracted data from DHIS2 in Microsoft Excel format were imported into R version 4.5.0 for cleaning, coding, and analysis. Data quality checks were performed by screening for missing values, duplicate entries, outliers, and inconsistencies. Inconsistent entries, outliers, and those with missing values of more than 5% were excluded from the final analysis. Among duplicate records, entries with more complete variable information were retained and used to populate missing data in less complete duplicates, after which the duplicate entries were dropped.

Descriptive analysis was done using frequencies and percentages, with SSI rates computed and expressed as the number of SSI cases per 100 surgical procedures performed in a given year. The trend of SSI across facilities was presented using line graphs, while variation in SSI rates across time in facility levels and facility ownership, and SSI type were assessed by linear regression using the Ordinary Least Squares method, and the significance of variations was measured at a 95% CI interval at p-value ≤ 0.05.

### Ethics consideration

Ethical approval was obtained from the National Health Research Ethics Committee (NatHREC) under certificate number Ref. NIMR/HQ/R.8a/Vol.IX/505, and additional authorization to use the DHIS2 data was obtained from the Tanzania Ministry of Health.

## Results

### Demographic characteristics of Health facilities reporting SSI data

Of 138 health facilities included in the final analysis, the majority were publicly owned, 86.2% (119/138), while 13.8% (19/138) were of private ownership, with 69.6% (96/138) being from the primary level, 22.5% (31/138) being of secondary level while 8% (11/138) being of tertiary level. Among 419,412 surgical procedures reported from health facilities in the DHIS2 database, 38.3% (160,501/419,412) were non-caesarean procedures, while 61.7% (258,911/419,412) were caesarean procedures. Table 1 shows the demographic characteristics of the study facilities.

**Table 1:** Demographic characteristics of Health facilities reporting SSI data from 2021 to 2025.

| Variables | N (%) |
| --- | --- |
| **Number of Health Facilities N=138** |  |
| Tertiary | 11(8) |
| Secondary | 31(22.5) |
| Primary | 96(69.6) |
| **Facility Ownership N=138** |  |
| Public | 119 (86.2) |
| Private | 19 (13.8) |
| **Type of Surgical Procedure N=419,412** |  |
| Non-C/S procedures | 160,501(38.3) |
| C/S procedures | 258,911(61.7) |

### Temporal national trends of SSI across Tanzania in health facilities

Despite an increase in facilities reporting SSI surveillance from 17 in 2021 to 138 in 2025, the national trend of surgical procedures has increased from 9,616 to 127,039 between 2021 and 2025, following the expansion of facilities reporting SSI rates from 17 in 2021 to 138 in 2025. During the same period, the rate of SSI cases decreased from 3.1% to 2.4%, with a sharp increase to 3.7% in 2022 (Figure 3).

**Fig 3:**
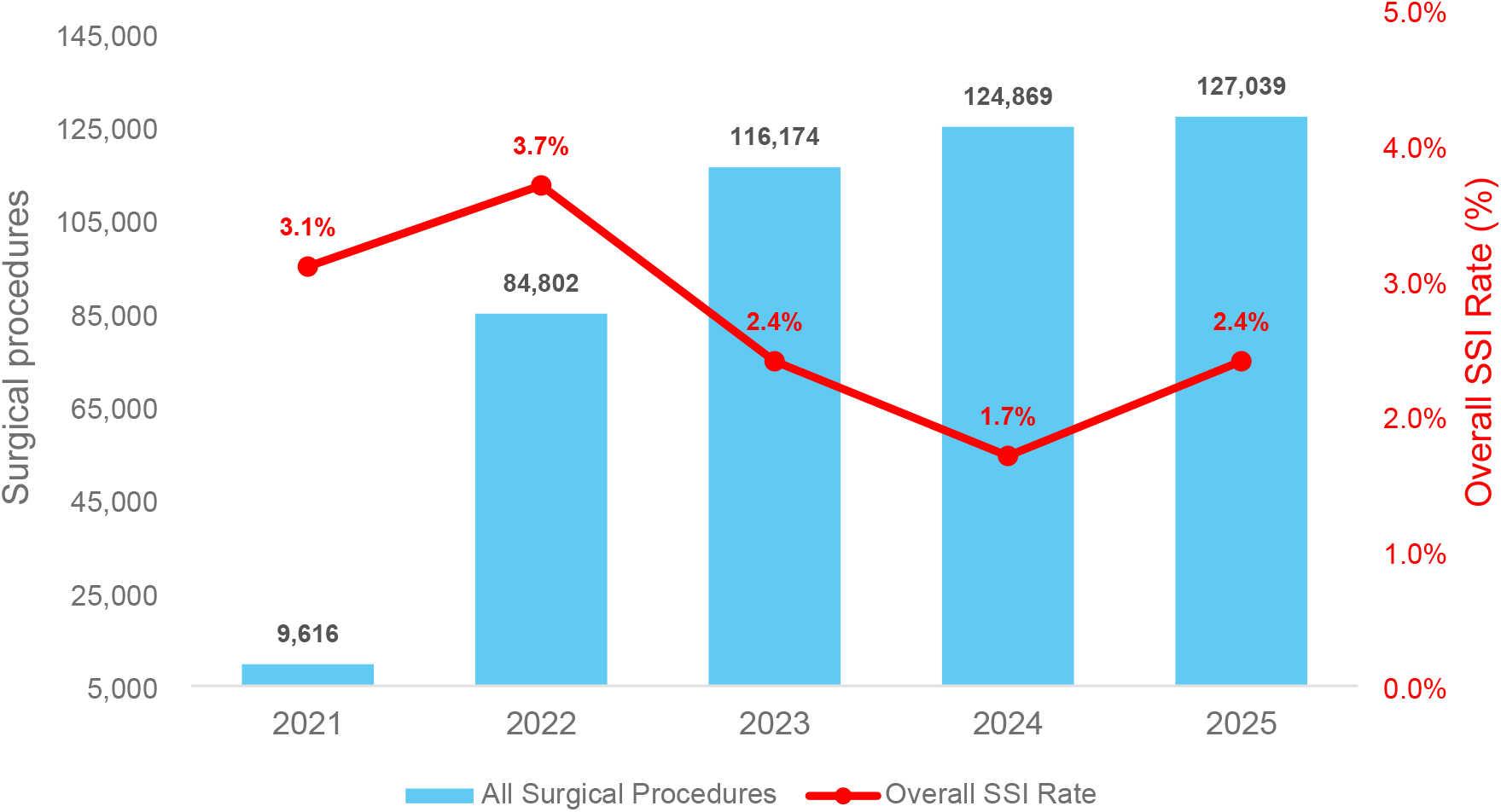
National trend of surgical procedures and SSI rate in Tanzania health facilities, 2021 - 2025

### The overall summary of Surgical procedures and SSI within health facilities from 2021 to 2025

Between 2021 and 2025, the number of reporting facilities increased substantially from 17 to 138. This expansion coincided with a marked increase in surgical procedures for both types, with non-caesarean procedures rising from 5715 to 48,300 and caesarean section procedures from 3,901 to 78,739. Over the same period, non-C/S SSI rates decreased from 3.3% to 1.3%, with a sharp increase to 5.1% in 2022, while the rates of C/S SSI remained relatively stable, with a slight increase from 1.8% to 2% from 2021 to 2025, as illustrated in Table 2.

**Table 2:** The overall summary of surgical procedures and SSI from 2021 to 2025.

| Year | Reporting Facilities | Non-CS Procedures | Non-CS SSI Cases N (%) | C/S Procedures | C/S SSI Cases N (%) |
| --- | --- | --- | --- | --- | --- |
| 2021 | 17 | 5,715 | 190(3.3) | 3,901 | 71.0 (1.8) |
| 2022 | 67 | 32,746 | 1,670(5.1) | 52,056 | 1,094(2.1) |
| 2023 | 110 | 44,259 | 885(2) | 71,915 | 1,295(1.8) |
| 2024 | 131 | 51,025 | 459(0.9) | 73,843 | 1,108(1.5) |
| 2025 | 138 | 48,300 | 628(1.3) | 78,739 | 157(2) |

### The rate of SSI by facility type and ownership

Between 2021 and 2025, the primary level facilities accounted for 5.4% SSI rates, higher compared to 1.6% and 1.7% observed in secondary and tertiary level facilities, respectively. Within the primary level facilities, SSI rates were higher in privately owned facilities (5.4%) compared to public-owned facilities (3.6%). Contrarily, at the tertiary level, SSI rates in public facilities were higher (1.7%) compared to 0.9% observed in privately owned facilities. At secondary level facilities, SSI rateS remained constant at 1.6% regardless of ownership (Figure 4).

**Fig 4:**
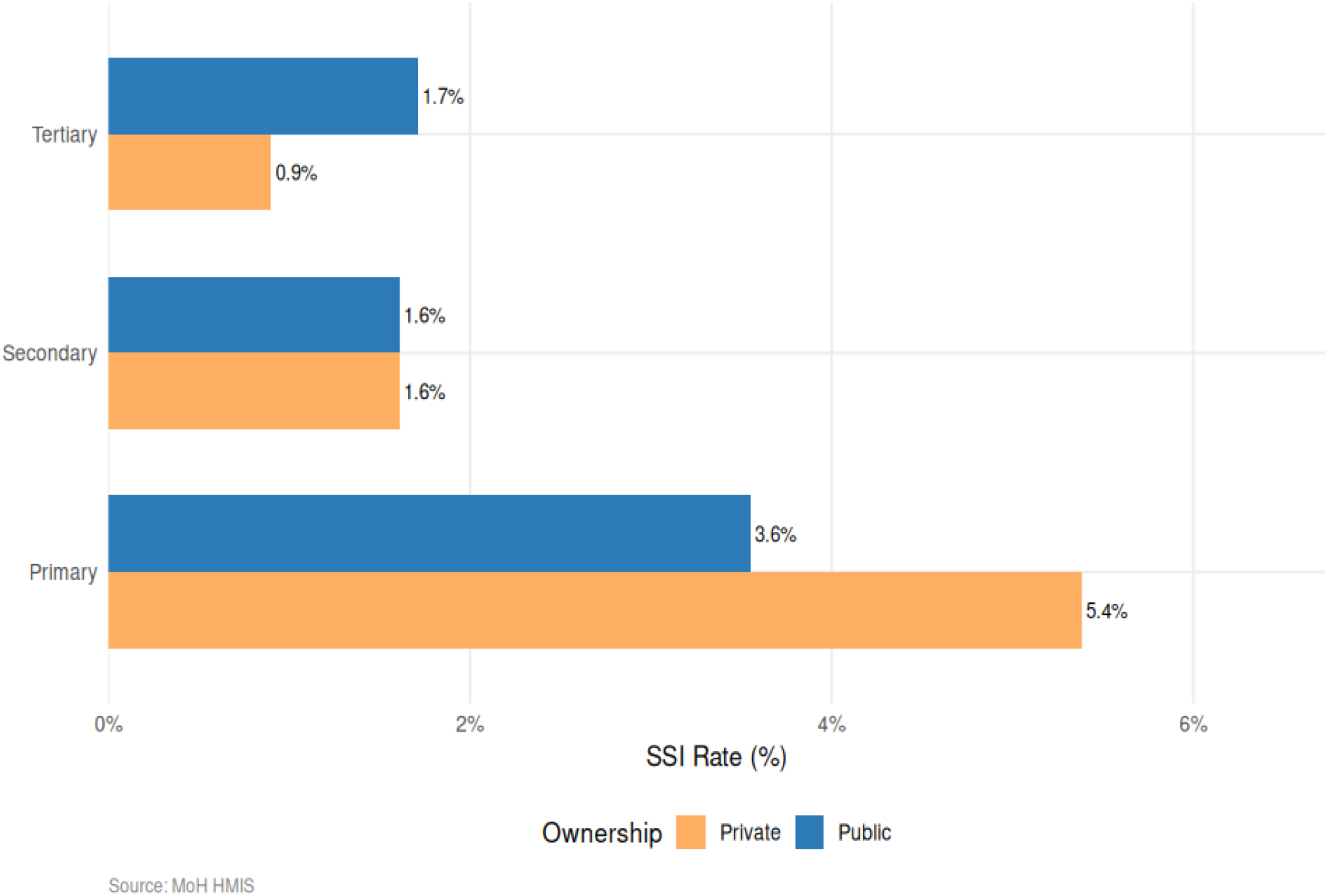
The pooled rate of SSI rates by facility type and ownership, 2021-2025

### Trend analysis of SSI across Tanzanian Health facilities between 2021 and 2025

Between 2021 and 2025, the Ordinary Least Squares (OLS) trend analysis showed overall SSI rates declined from 3.1% to 2.4%, though this trend was not statistically significant. The rates of SSI in primary-level facilities were consistently higher, between 4.7% to 4.0%, compared to the tertiary-level hospitals, where SSI rates were between 1% and 1.5%. No statistically significant trends were observed by facility level or ownership.

Among the SSI types, there was no significant change in infection rate for type 1(superficial) and type 2(deep) surgical site infections. However, type 3 (organ/space) infections demonstrated a statistically significant decrease, with rates declining from 1.2% to 0.3%, (-0.23%/year, 95% CI: -0.34 to -0.12; p=0.006), illustrated in Table 3.

**Table 3:**
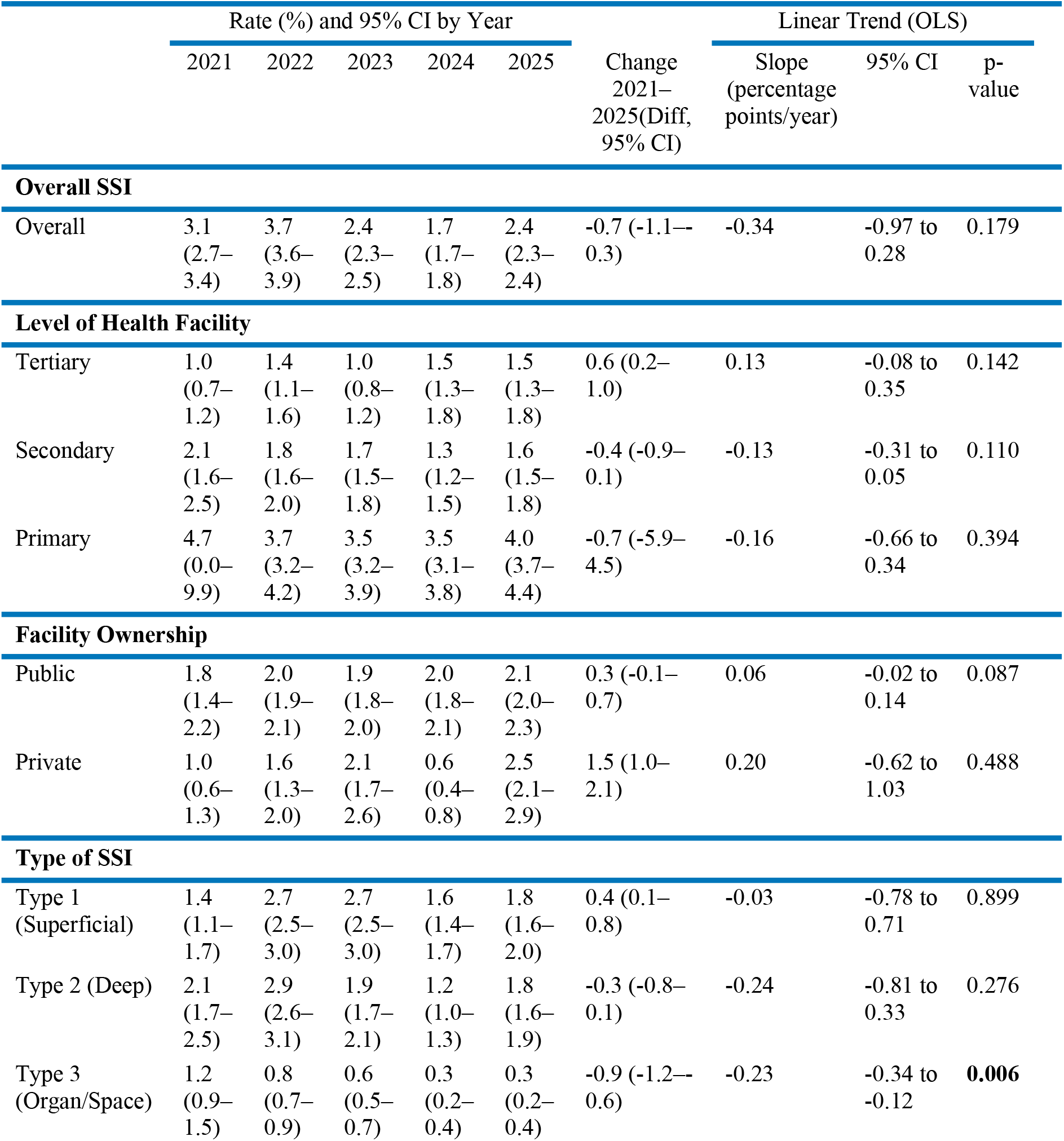
Trend analysis of SSI rates by facility-level, facility ownership, and type of procedure between 2021 and 2025.

## Discussion

The current study has attempted to estimate the burden of SSI by determining not only trend rates but also SSI rates across different levels of health facilities, and the variation of SSI rates by SSI type, health facility level, and facility ownership in Tanzania.

The study findings indicated that the SSI rate declined from 3.1% to 2.4%, despite increased surgical volume. The SSI rates described in this study aligned with the pooled SSI incidence rate of 2.5% reported by Dechassa Adere Mengistu et al. ^26^. In contrast, the result of the current study is lower than the rates of 5% and 4% reported in India and China, and in other SSA countries, such as Ethiopia (12.3%), Nigeria (5.1%), and Uganda (6%) ^27,28^. The observed overall lower SSI rate found in the current study likely reflects the effectiveness of ongoing SSI surveillance implementation across the study facilities in the country^29^. Imanishimwe et al found that participation in SSI surveillance networks alone was associated with a 35% reduction in SSI rates over nine years across 17 high-income country networks^20^. There is a need to scale up integrated SSI prevention strategies, such as combining surveillance, multimodal IPC interventions, and capacity strengthening as a cost-effective pathway to achieving sustained reductions in surgical morbidity across all Tanzanian health facilities and similar settings.

The rate of SSI due to caesarean section in the current study relatively increased from 1.8% to 2% over the study period, aligning with global country-level patterns where pooled incidence estimates have steadily increased over time, with an incidence rate of 5.6% reported in 2023 ^26^. Contrarily, the findings are substantially lower than the regional average of 11.9% across African countries ^30^. This difference may be attributed to the study’s geographical location and study designs, as previous studies were conducted in a pool of many countries with different grounds of health policies, using a meta-analysis approach, while the current study enrolled a few eligible facilities from Tanzania using a cross-sectional study design. Despite the low absolute rate, a significant increase in the SSI trend observed in this study confirms an increased SSI burden among women who underwent C-sections, posing a potential risk to maternal morbidity, mortality, repeated surgeries, and socioeconomic effects ^31^. Thus, this situation underscores the necessity of strengthening effective IPC protocols in obstetric care.

The current study found that the rate of SSI was highest in primary level facilities compared to the tertiary level facilities. This might be due to poor adherence to IPC practices in health centers and dispensaries, as reported earlier by Kinyenje et al ^32^. However, other factors like a shortage of essential health commodities, including proper antiseptic solutions and sterile materials, as well as inappropriate use of antibiotics, have been reported to contribute to an increased rate of SSI in primary health facilities^33^.

Furthermore, the study findings revealed that high rates of SSI are occurring in public hospitals (1.7%) compared to private hospitals (0.9%) at the tertiary level, unlike at the primary level, where higher rates of SSI were more reported in privately owned facilities. This might be due to the low proportion (13.8%) of private hospitals in the study. These findings align with an Ethiopian study that found higher SSI rates (13.4%) in public hospitals compared to private hospitals (6.5%) ^34^. The plausible explanation for the observed differences could be due to a systemic challenge of higher patient-to-staff ratios in public settings compared to private ones, considering that tertiary levels are the referral centres that serve primary and secondary levels in LMICs health systems, thus operating in substandard conditions with poor IPC practices. Moreover, private hospitals are relatively clean and implement strict IPC strategies to increase patient satisfaction. Contrarily, C. Schroder et al found lower SSI rates in public hospitals following hip procedures in Germany ^35^. This might be due to differences in health system organisational structure, as German public hospitals operate under mandatory national surveillance systems with increased specialization and experience to maintain high-quality IPC.

Considering the SSI type, the organ/space infections decreased significantly from 1.2% in 2021 to 0.3% in 2025, unlike superficial and deep SSI, which showed no statistically significant changes. Given that organ/space SSI is often associated with more severe clinical outcomes, prolonged hospitalization, and increased healthcare costs, these findings indicate meaningful progress in quality and patient safety ^36^. Several factors may have contributed to this decline, including the gradual strengthening of SSI surveillance systems, improved adherence to IPC, and increased awareness of perioperative risk reduction strategies. Strengthening standard operating procedures, including routine change of gloves and instruments before abdominal wound closure, is of paramount importance and has been proven to reduce 13% of SSI rates by 30 days after surgery across a wide range of hospitals, including both large, tertiary-level hospitals with advanced perioperative services and even small, rural hospitals with fewer resources ^37^. However, the gradual increase in superficial SSI rates suggests the need for uniform efforts to strengthen compliance with evidence-based SSI prevention measures. Sustained implementation of WHO recommendations on SSI prevention, including surveillance, environmental hygiene in the operating room, and the decontamination of medical devices and surgical instruments, remains critical to achieve further reduction of SSI burden ^26^.

### Strength and Limitations of this study

To what extent our findings are generalizable to similar settings remains to be seen; however, our results do offer an idea of the dimension of SSIs across Tanzanian Health facilities and highlight the importance of targeted IPC interventions. Additionally, the use of routinely collected surveillance data from the national aggregated DHIS2 database reflects real-world clinical practice and strengthens the public health relevance of the findings.

However, the analysis relied on secondary routine health facility data; the findings may be affected by underreporting and reporting inconsistencies across facilities. Also, the study lacked patient-level clinical information such as comorbidities, duration of surgery, antimicrobial use, and postoperative follow-up, preventing adjustment for confounding factors. Lastly, being a cross-sectional study makes it difficult to infer causality. Although these limitations may have introduced some bias or reduced the study’s generalizability, their overall impact on the study’s conclusions is considered minimal.

## Conclusion and recommendation

This study aimed to evaluate trends and variation in SSI rates in Tanzanian health facilities. To our knowledge, this is the first nationwide study to evaluate the trends and variation of SSI rates using a large number of observations delivered from monthly facility-level SSI reporting over five years.

The study found that overall SSI rates in Tanzanian health facilities relatively declined despite increasing surgical volume. However, SSI rates following the caesarean section increased over time, with higher rates observed in primary and public health facilities. Although trend analysis showed no significant change in infection rates for type 1(superficial) and type 2(deep) SSI, type 3 (organ/space) infections demonstrated a statistically significant decline. These findings suggest persistent disparities in infection prevention and control capacity, availability of essential resources, and implementation of evidence-based surgical safety practices across facility levels and ownership types.

These findings highlight the need to strengthen SSI surveillance systems and ensure effective translation of the quality improvement interventions into routine surgical practice. Particular emphasis should be placed on adherence to WHO-recommended perioperative care bundles, antimicrobial prophylaxis, and surgical safety practices, with focused support for obstetric care in primary and public health facilities to further reduce preventable SSI and improve the quality and safety of surgical care in Tanzania and similar settings

## Acknowledgements

We firmly acknowledge the technical support from the Quality Assurance Unit at the MoH, Prime Minister’s Office, Regional Administration and Local Government (PMO-RALG), the University of Dar es Salaam DHIS2 Unit, and the Global Health Security project implemented by the Centre for International Health, Education, and Biosecurity (CIHEB) Tanzania. We also extend our sincere appreciation to all healthcare workers involved in routine collection and reporting of data through the District Health Information System (DHIS2).

## List of observations

AMR: Antimicrobial Resistance
DHIS2: District Health Information System
HAIs: Hospital-Acquired Infections
HH: Hand Hygiene
IPC: Infection Prevention and Control
SSI: Surgical Site Infections

## Authors’ Contributions

**Conceptualization:** Cesilia Charles, Jonhas Masatu, Melkizeseki Abdulahi, Elford Mukerebe, and Erick Kiyenje

**Data curation:** Upendo Msanjila, Nuru Beda, Christina Mmasa, Adam Kamese

**Formal analysis:** Upendo Msanjila and Adam Kamese

**Investigation:** Edgar Lusaya, Joseph C Hokororo, and Eliudi S. Eliakimu

**Methodology:** Furahini Mbise, Calvin Andrew, Omary Nassoro, and Lackson David

**Project administration:** Edgar Lusaya, Joseph C Hokororo, and Eliudi S. Eliakimu

**Resources:** Siril Kullaya, Edgar Lusaya, Joseph C Hokororo, and Eliudi S. Eliakimu

**Software:** Upendo Msanjila and Adam Kamese

**Supervision:** Siril Kullaya, Edgar Lusaya, Joseph C Hokororo, and Eliudi S. Eliakimu

**Validation:** Radenta Bahegwa and Laura Marandu

**Writing original draft:** Cesilia Charles, Jonhas Masatu, Elford Mukerebe, and Erick Kiyenje

**Writing review and editing:** Cesilia Charles, Erick Kiyenje, Christina Mmasa, Furahini Mbise, Joseph C Hokororo, and Eliudi S. Eliakimu

## Artificial Intelligence disclosure

Artificial intelligence was not utilized in any aspect of manuscript preparation, including data analysis and interpretation. However, on select occasions, it was employed solely to identify and correct grammatical errors

## Consent for publication

All authors consented to publish this study

## Provenance and peer review

Not commissioned; not externally peer reviewed

## Data availability statement

All data are available upon reasonable request via

## Disclaimer

The Ministry of Health, as well as all stakeholders who implemented IPC M&E, had no role in the writing of the article. The findings and conclusions contained within are those of the authors and do not necessarily reflect the positions or policies of the Ministry and other stakeholders.

